# Effects of a bone-strengthening exercise intervention on bone health in pediatric cancer survivors: A Randomized Controlled Trial

**DOI:** 10.1101/2025.04.21.25325629

**Authors:** Andres Marmol-Perez, Esther Ubago-Guisado, Jose J. Gil-Cosano, Andrea Rodriguez-Solana, Francisco B Ortega, Maria Elena Mateos Gonzalez, Juan Francisco Pascual-Gazquez, Francisco J Llorente-Cantarero, Cristina Cadenas-Sanchez, Vicente Martinez-Vizcaino, Kirsten K Ness, Jonatan R Ruiz, Luis Gracia-Marco

**Author notes:** Corresponding author Esther Ubago-Guisado.

## Abstract

Pediatric cancer survivors remain at increased risk of low areal bone mineral density (aBMD) after treatment. We investigated the effects of a 9-month online bone-strengthening exercise intervention on the femoral neck aBMD Z-score (primary outcome), and other bone health parameters measured using dual-energy X-ray absorptiometry at the hip, total body, and lumbar spine in young pediatric cancer survivors. A total of 116 participants were randomized into the exercise or control groups (N = 58 in each). The exercise group performed a 9-month periodized video-recorded bone-strengthening exercise intervention based on squats and jumps (3–4 days/week for 10–20 min/session). No between-group differences in femoral neck aBMD Z-score (Cohen’s d = −0.08, *P* = .706) and aBMD Z-score outcomes were observed post-intervention. However, the intervention showed a small effect size on the total hip, with a borderline non-significant improvement in aBMD Z-score (Cohen’s d = 0.35, *P* = .054) and a significant improvement in BMC Z-score (Cohen’s d = 0.38, *P* = .039). No adverse events were reported. Adherence was high (87.4%). The 9-month online bone-strengthening exercise intervention did not increase the femoral neck aBMD Z-score; however, it improved the overall BMC Z-score at the hip in young pediatric cancer survivors. This intervention is safe, well-tolerated, and may contribute to reduce the risk of bone impairments during adulthood.

**Trial Registration:** isrctn61195625 dated April 2, 2020.

**One Sentence Summary:** A 9-month online bone-strengthening exercise intervention does not improve bone health at the femoral neck in pediatric cancer survivors.

## INTRODUCTION

Cancer treatments in early life increase the risk of long-term health complications,(*1*) including low areal bone mineral density (aBMD),(*2*) prevalent in approximately two-thirds of survivors.(*3*) Non-pharmacological treatments, including exercise,(*4*) improve aBMD in healthy children if exercise training principles (i.e., frequency, intensity, time, type, volume, and progression) are appropriately followed. However, randomized controlled trials (RCTs) in this population(*5*) targeting bone outcomes such as femoral neck aBMD, a key outcome in the diagnosis of osteoporosis during adulthood,(*6*) are limited.

Prior exercise interventions in young pediatric cancer survivors have been ineffective,(*5*) which could be attributed to the short duration of trials (3 months),(*7*) inclusion of non-osteogenic exercises (aerobic exercise),(*8*) or microgravity environment (swimming pools).(*7*) Moreover, meeting the exercise frequency and intensity required to improve bone outcomes is challenging for children during treatment.(*9*) Therefore, exercise-based RCTs specifically designed to improve bone health after pediatric cancer are required. This randomized controlled trial aimed to investigate the effects of a 9-month online bone-strengthening exercise intervention on femoral neck aBMD Z-scores (primary outcome) and other bone health parameters (secondary outcomes) in young pediatric cancer survivors. We hypothesized that this intervention improves bone health in this population.

## RESULTS

### Study participants

Among the 196 invited participants, 116 provided consent (recruitment rate: 59.2%) (Fig. 1). In the exercise group, 13 participants did not complete the post-intervention bone health evaluation and nine did not meet the adherence requirements. In the control group, four participants did not complete the post-intervention bone health evaluation. The median (interquartile range) adherence in the exercise group was 93.8% (9.4), 90.9% (19.3), and 73.3% (34.2) in the first, second, and third phases, respectively, resulting in a total adherence rate of 87.4% (12.7). All 116 participants were included in the intention-to-treat analysis, and 90 (77.6%) were included in the planned per-protocol analysis. No significant differences, except for radiotherapy exposure, were observed between dropouts and non-dropouts (*P* = .014) (Supplementary Table 2).

**Fig. 1.**
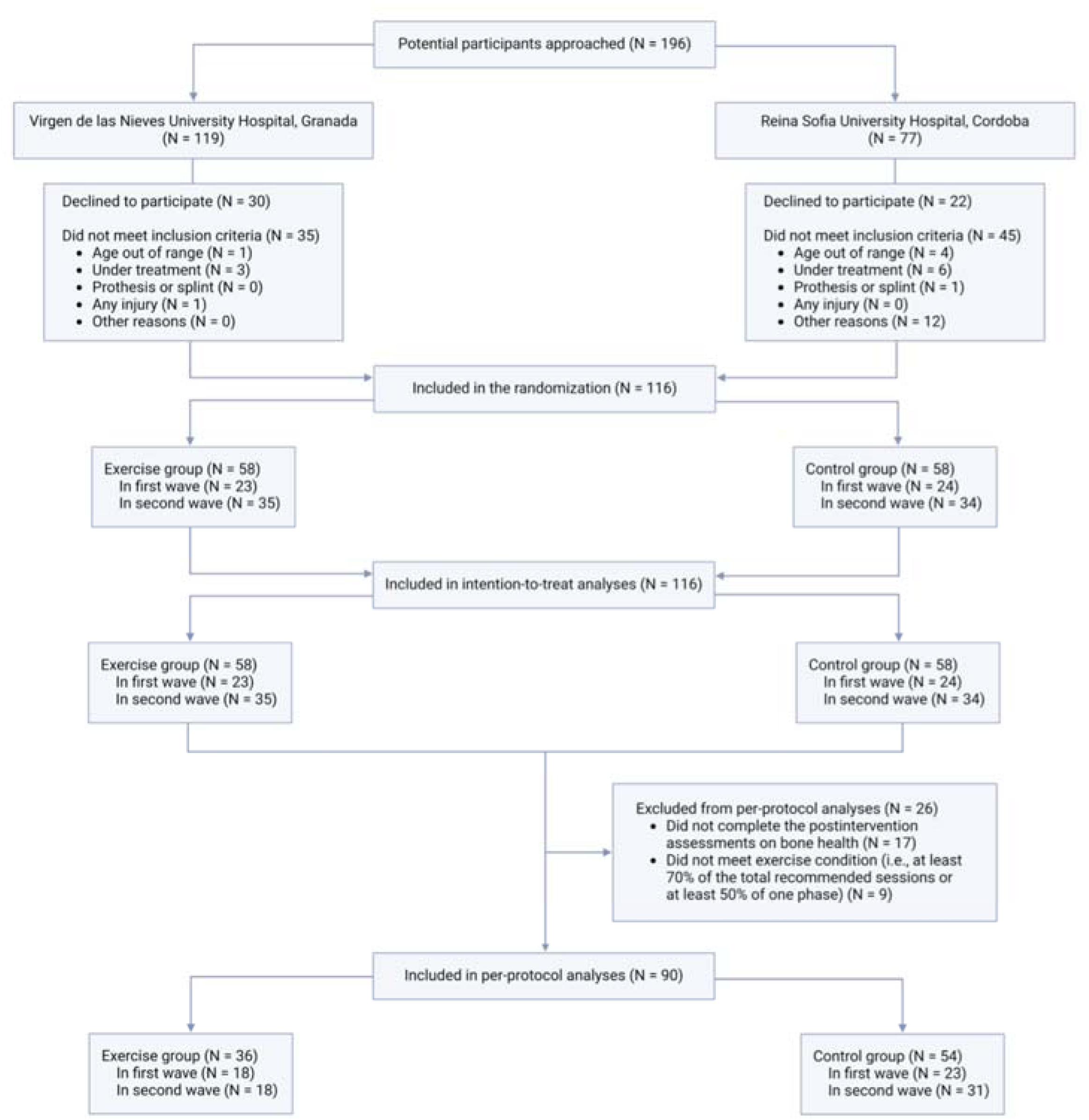
Flow chart of participant selection.

The baseline characteristics of the participants are listed in Table 1. Of the included participants, 58 were randomized to the exercise group (mean [SD] age: 11.7 [3.2] years [range 6.4%–17.7]; 37.9% were girls) and 58 were randomized to the control group (mean [SD] age: 12.5 [3.5] years [range: 6.4–18.0]; 46.6% were girls). The proportion of participants exposed to radiotherapy was comparable between the groups (27.6%). The aBMD and BMC Z-score were slightly higher in the exercise group than in the control group for all outcomes. Acute lymphoblastic leukemia was the most common cancer type (39.7% and 37.9% in the exercise and control groups, respectively [Supplementary Table 3]).

**Table 1.**
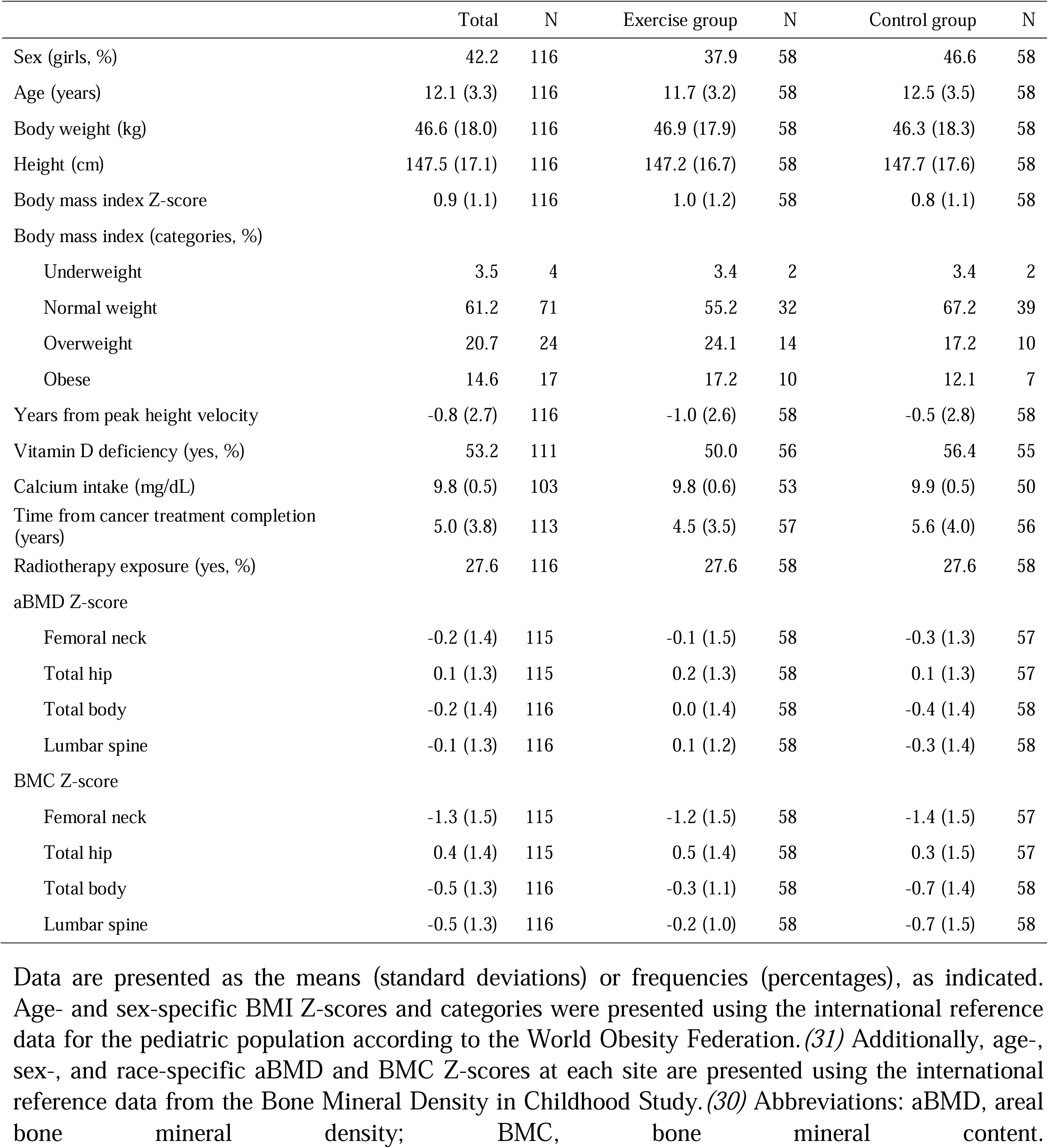
Baseline characteristics of the included participants.

### Changes in aBMD and BMC Z-scores

Table 2 shows within- and between-group differences in primary and secondary outcomes from baseline to end of intervention. No between-group differences were observed in femoral neck aBMD Z-score (Cohen’s d = - 0.08, *P* = .706) or other aBMD Z-score outcomes. However, the intervention had a small-sized borderline non-significant effect on total hip aBMD Z-score (Cohen’s d = 0.35, *P* = .054) and a significant effect on total hip BMC Z-score (Cohen’s d = 0.38, *P* = .039). No other between-group differences were observed. Compared with the control group, the exercise group demonstrated small-sized effects ranging from 0.26 to 0.28 (*P* = .168 to .124) for the femoral neck, total body, and lumbar spine BMC Z-scores. Estimated means at constrained baseline and 9-month follow-up in the aBMD and BMC Z-score outcomes are presented in Fig. 2. Subgroup analyses (available in the supplementary online materials, Fig. S1) revealed descriptive findings based on the interactions of age (lumbar spine BMC Z-score), sex (femoral neck, total hip, total body, and lumbar spine BMC Z-score), somatic maturity (lumbar spine BMC Z-score), and bone health baseline values (total hip BMC Z-score). These effects remained consistent after multiple-testing correction, except for total hip BMC Z-score (adjusted *P* = .216).

**Fig. 2.**
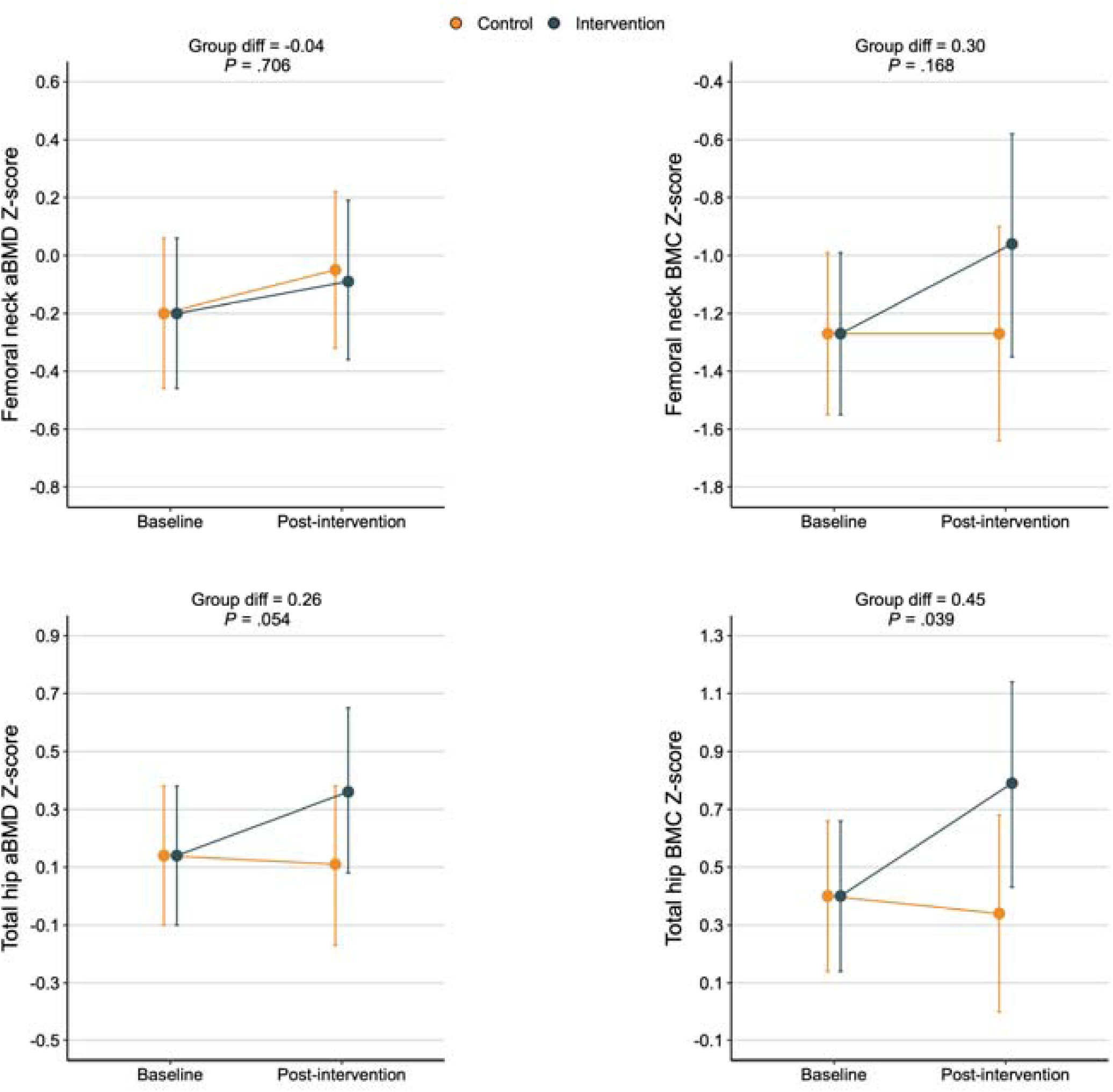
Estimated means at constrained baseline and 9-month post-intervention follow-up in femoral neck and total hip aBMD and BMC Z-score outcomes. Error bars represent 95% confidence intervals. Data were analyzed using a constrained longitudinal data analysis (cLDA) with linear outcomes, assuming baseline means of the outcome measure were identical between the groups (intention-to-treat analysis). Abbreviations: aBMD, areal bone mineral density; BMC, bone mineral content; Diff, difference.

**Table 2.** Baseline, within-group changes (baseline to 9-month follow-up) and between-group differences (exercise vs. control) in the aBMD and BMC Z-score outcomes (intention-to-treat analysis)

|  | Exercise group |  | Control group |  | Between-group differences |  |  |
| --- | --- | --- | --- | --- | --- | --- | --- |
|  | Mean (95% CI) | Change %<sup>Ψ</sup> | Mean (95% CI) | Change %<sup>Ψ</sup> | MD (95% CI) | Effect size | P<sup>¶</sup> |
| Femoral neck aBMD Z-score |  |  |  |  |  |  |  |
| Baseline | -0.20 (-0.46 to 0.06) | NA | -0.20 (-0.46 to 0.06) | NA | NA | NA | NA |
| Changes at 9-month | 0.11 (-0.05 to 0.28) | 4.11 | 0.15 (0.00 to 0.30) | 5.48 | -0.04 (-0.22 to 0.15) | -0.08 | .706 |
| Total hip aBMD Z-score |  |  |  |  |  |  |  |
| Baseline | 0.14 (-0.10 to 0.38) | NA | 0.14 (-0.10 to 0.38) | NA | NA | NA | NA |
| Change at 9-month | 0.22 (-0.01 to 0.46) | 6.10 | -0.03 (-0.25 to 0.18) | -3.66 | 0.26 (0.00 to 0.52) | 0.35 | .054 |
| Total body aBMD Z-score |  |  |  |  |  |  |  |
| Baseline | -0.19 (-0.44 to 0.06) | NA | -0.19 (-0.44 to 0.06) | NA | NA | NA | NA |
| Changes at 9-month | 0.03 (-0.16 to 0.23) | 3.80 | -0.06 (-0.24 to 0.12) | -3.80 | 0.09 (-0.13 to 0.31) | 0.15 | .422 |
| Lumbar spine aBMD Z-score |  |  |  |  |  |  |  |
| Baseline | -0.07 (-0.32 to 0.18) | NA | -0.07 (-0.32 to 0.18) | NA | NA | NA | NA |
| Changes at 9-month | -0.05 (-0.19 to 0.08) | -4.11 | -0.02 (-0.14 to 0.10) | -4.11 | -0.04 (-0.19 to 0.12) | -0.07 | .642 |
| Femoral neck BMC Z-score |  |  |  |  |  |  |  |
| Baseline | -1.27 (-1.55 to -0.99) | NA | -1.27 (-1.55 to -0.99) | NA | NA | NA | NA |
| Changes at 9-month | 0.30 (-0.08 to 0.69) | 10.51 | 0.00 (-0.36 to 0.36) | 7.25 | 0.30 (-0.13 to 0.73) | 0.26 | .168 |
| Total hip BMC Z-score |  |  |  |  |  |  |  |
| Baseline | 0.40 (0.14 to 0.66) | NA | 0.40 (0.14 to 0.66) | NA | NA | NA | NA |
| Changes at 9-month | 0.39 (0.00 to 0.77) | 14.83 | -0.06 (-0.42 to 0.30) | -8.66 | 0.45 (0.02 to 0.87) | 0.38 | **.039** |
| Total body BMC Z-score |  |  |  |  |  |  |  |
| Baseline | -0.49 (-0.73 to -0.25) | NA | -0.49 (-0.73 to -0.25) | NA | NA | NA | NA |
| Changes at 9-month | 0.10 (-0.07 to 0.27) | 8.52 | -0.04 (-0.20 to 0.12) | -7.08 | 0.14 (-0.06 to 0.33) | 0.26 | .171 |
| Lumbar spine BMC Z-score |  |  |  |  |  |  |  |
| Baseline | -0.45 (-0.68 to -0.21) | NA | -0.45 (-0.68 to -0.21) | NA | NA | NA | NA |
| Changes at 9-month | 0.11 (-0.06 to 0.28) | 9.21 | -0.04 (-0.19 to 0.12) | -7.07 | 0.15 (-0.04 to 0.34) | 0.28 | .124 |

The results are presented as the mean change from baseline for each group and the mean difference between the groups. Data were analyzed using constrained longitudinal data analysis (cLDA) with linear outcomes, assuming that t baseline means of the outcome measure were identical between the groups. Age-, sex-, and race-specific aBMD and BMC Z-scores at each site are presented using the international reference data from the Bone Mineral Density in Childhood Study.(30) Effect sizes were calculated using the Cohen's d formula. Abbreviations: aBMD, areal bone mineral density; BMC, bone mineral content; CI, confidence interval; MD, mean difference; NA, not applicable.
<sup>‡</sup> The percentage change (9-month follow-up minus baseline values/baseline values × 100) was calculated from the raw values (g/cm<sup>2</sup> or g).
□ P-values indicate between-group indicate between-group differences based on the aBMD and BMC Z-score outcomes.

In the per-protocol analysis, the findings for aBMD Z-score outcomes were consistent with those from the intention-to-treat analyses (Supplementary Table 4). However, the effect sizes of the intervention on BMC outcomes increased in the per-protocol than in the intention-to-treat analyses (Cohen’s d = 0.42 to 0.43, *P* = .050 to .043) for the femoral neck, total body and lumbar spine, and remained similar (Cohen’s d = 0.39, *P* = .057) for the total hip. Using this approach, the estimated means at constrained baseline and 9-month follow-up in the aBMD Z-score outcomes and raw individual changes in the total hip and femoral neck aBMD Z-scores from baseline to 9-month follow-up were analyzed (Fig. S2). No adverse events were reported.

### Adverse Events

There were no reported adverse events regarding the intervention.

## DISCUSSION

Our findings indicated that the 9-month online bone-strengthening exercise intervention did not significantly increase the femoral neck aBMD Z-score; however, it improved the overall BMC Z-score at the hip in young pediatric cancer survivors. Notably, among those who completed the minimum recommended number of exercise sessions (per-protocol analysis), nearly medium-sized effects were observed for the femoral neck BMC Z-score.

Contrary to a previous systematic review of healthy children and adolescents that reported beneficial effects of plyometric exercise-based interventions on femoral neck aBMD,(*4*) our intervention was not effective in this regard. Although unsupervised exercise interventions improve bone health in adult populations,(*10*) the lack of supervision in our study involving growing children may have affected its effectiveness. In particular, supervision can ensure real compliance, continuous engagement, immediate feedback, and motivation in this population. However, previous non-osteogenic exercise interventions in young pediatric cancer survivors(*7, 8*) demonstrated similar non-significant effects on femoral neck aBMD Z-scores and aBMD, suggesting that this specific region could be less responsive.

The small-sized effects on the total hip and femoral neck BMC Z-scores (intention-to-treat and per-protocol analyses, respectively) in our study were consistent with those observed in previous meta-analyses on BMC outcomes in healthy children and adolescents; (*11, 12*) however, similar results were also found for aBMD outcomes. A recent similar meta-analysis in this population reported greater increases in BMC outcomes (standardized mean difference [SMD] = 1.348; 95% CI, 1.053–1.643) than in aBMD outcomes (0.640; 95% CI, 0.417–0.862).(*13*) BMC (g) is a measure of the amount of mineral present in the bone, and aBMD (g/cm^2^) is a volumetric measure that accounts for both mineral quantity in grams and the area of the bone assessed by DXA. The optimal intervention duration to improve bone health in previous studies ranged from 8 to 12 months(*4*), aligning with our study duration. In addition, the inclusion of behavioral change techniques, as in our intervention, is recommended,(*14, 15*) which likely contributed to the high adherence rates (83%) among our study participants. Improvements observed during childhood may have important public health implications, such as reduction in osteopenia/osteoporosis risk,(*16, 17*) subsequent fractures,(*18*) and the need for hospitalization, rehabilitation, or future disability in adulthood.(*2*)

Although our bone-strengthening exercise intervention was not effective in improving bone health in the total body, small-sized effects were observed on total-body BMC Z-scores (per-protocol analysis). Plyometric exercises improved total body BMD and BMC outcomes in healthy children and adolescents.(*4*) Mogil et al.(*19*) observed substantial improvements in total body aBMD Z-scores in young pediatric cancer survivors following a low-magnitude, high-frequency mechanical stimulation for 1 year. This intervention was conducted 7 days/week, with 2 sessions per day, each lasting 10 min. The positive outcomes may be attributed to the frequency (twice per day) and adequate intervention duration (1 year). Consistent with our findings, a non-RCT evaluating the effects of a concurrent full-body exercise intervention for 6 months (3 days/week, 55–60 min per session) in survivors did not report significant effects on total body aBMD Z-scores.(*8*) Although our intervention was specifically designed to improve bone health following the training principles employed in previous successful interventions in healthy children and adolescents,(*20, 21*) it lacked core and upper-body exercises that might have simulated muscle growth or activation and associated bone remodeling.(*22*) This could explain the small-sized effects (borderline) observed on lumbar spine BMC Z-scores (per-protocol analysis). Altogether, our findings suggest that bone adaptations may be site-specific because observed effects were mainly found at the hip region, which was our main target.

Our study had certain limitations, which should be considered while interpreting the results of this RCT. First, although adherence to the intervention was monitored using a monthly diary review, the exercise sessions were not supervised throughout the intervention. Nevertheless, parents provided necessary support when needed, and the research staff maintained frequent contact via chat groups. Notably, we observed higher positive effects among those who completed a minimum number of exercise sessions (per-protocol analysis). Second, COVID-19 restrictions may have affected the efficacy of this RCT; however, we lack data to confirm this in our sample. Third, recruiting participants was challenging due to the rarity of pediatric cancer. Future studies should consider evaluating the benefits of a similar bone-strengthening exercise intervention in survivors with low aBMD. Fourth, missing values at 9-month post-intervention follow-up were considerably higher in the exercise group than in the control group, yet the percentage of missing values were low and there were not significant differences between dropouts and non-dropouts, except for radiotherapy exposure.

## Conclusion

Although the 9-month online bone-strengthening exercise intervention did not increase the femoral neck aBMD Z-score, it improved the overall BMC Z-score at the hip in young pediatric cancer survivors. Future studies should focus on survivors with low aBMD and incorporate supervised full-body exercise sessions that engage both muscles and bones to stimulate bone remodeling, which can be effectively performed remotely using current technologies.

## METHODS

### Study Design

The REBOTA-Ex parallel-group RCT (part of the iBoneFIT project) was approved by the Ethics Committee on Human Research of the Regional Government of Andalusia (Reference: 4500, December 2019), followed the ethical guidelines of the Declaration of Helsinki (revised version 2013), and was registered on isrctn.com (Reference: isrctn61195625, April 2, 2020).(*23*) The study followed the consolidated standards of reporting trials (CONSORT) reporting guidelines (Supplementary Table 1). All research processes were conducted in accordance with the Singapore Statement on Research Integrity.(*24*)

### Study Participants

Pediatric cancer survivors aged 6–18 years, diagnosed at least 1 year prior to enrolment, who had received radiotherapy and/or chemotherapy but were not currently undergoing cancer treatments, were recruited from the Pediatric Oncology and Hematology units of the ‘Virgen de las Nieves’ (Granada) and ‘Reina Sofia’ (Cordoba) University Hospitals. Exclusion criteria included simultaneous participation in another study placing participants at any additional risk, discomfort or affect the results of both studies, previous diagnosed anorexia nervosa/bulimia, known pregnancy and/or known alcohol and drug abuse, children requiring chronic oral glucocorticoid therapy, having an injury that may affect daily life activities and can be aggravated by exercise and having a lower limb prosthesis that prevent bone assessment. Owing to COVID-19 restrictions, data collection was performed in two phases: 1) October 2020 to November 2021 and 2) December 2021 to January 2023. Sample size was calculated based on an expected effect size of 0.25 for the change in femoral neck aBMD (key outcome in the diagnosis of osteoporosis),(*6*) with an α level of 0.05 and power of 80%. Considering a 20% loss to follow-up and potential subgroup analysis by age, a minimum of 116 participants was required (N = 58 each in the exercise and control groups).

### Randomization and Blinding

Following baseline assessments, the participants were randomized (1:1) to the exercise or control group using SAS software (version 9.1, SAS Institute Inc.) by an external partner (VMV) who was not involved in the recruitment, enrolment, evaluation or exercise intervention. Preparation for this randomization was done prior to baseline assessments. While outcome assessors were blinded to group allocation, participants could not be blinded because of the nature of exercise intervention.

### Intervention

The exercise group performed periodized online bone-strengthening exercise interventions at home. Sessions were video-recorded and delivered to participants every 2 weeks and included a total of 7296 squats/jumps (2000 squats and 5296 jumps) in 136 sessions, each lasting for 10–20 min. The intensity progressively increased in volume (three to four sets, 10–20 repetitions, and three to four sessions per week) during the intervention phase (fixed protocol). The intervention was divided into three phases with different durations and impact loadings. Phase 1 corresponded to the first 8 weeks of the exercise intervention and participants performed body mass-based squats. Phase 2 lasted for 12 weeks, and the participants performed squat jumps. Phase 3, the longest phase, lasted 16 weeks and the participants performed countermovement jumps. Additionally, the exercise intervention incorporated five behavior change techniques and a gamification design (featuring points and rankings) to maintain participants’ engagement and adherence.(*25–27*)

Educational leaflets and infographics based on the current recommendations for calcium and vitamin D(*28*) intake were delivered to all participants before the intervention. The participants in the control group continued their usual routines and were offered the same online bone-strengthening exercise intervention at the end of the trial (waitlist control group).

### Adherence

The intervention was remotely monitored using a diary provided at baseline. The minimum adherence allowed during the first phase of the intervention without justified reasons was 50%; however, the overall adherence at 9 months was required to be 70% (completion of 95/136 sessions). If a participant did not complete 70% of the intervention at the end of the 9 months but was able to reach 70% within 2 additional weeks, the exercise intervention was extended. To encourage adherence, participants were contacted monthly by the research staff to review the number of sessions completed and to provide problem-solving strategies.

### Data Collection Outcomes

Primary (femoral neck aBMD [g/cm^2^]) and secondary outcomes (aBMD and bone mineral content [BMC, g]) were measured using dual-energy X-ray absorptiometry (DXA) at hip regions (femoral neck and total hip), total body (less head), and lumbar spine (mean of L1–L4) at baseline and 9-month follow-up. Scanning was performed using a single DXA scanner (Hologic Series Discovery QDR, Bedford, MA, USA) and analyzed using APEX software (version 4.0.2). The device was calibrated daily using a lumbar spine phantom. The participants were instructed to remain still in the supine position during scanning.(*6*) All DXA scans were analyzed by a licensed and experienced researcher. The coefficient of variation for DXA in the pediatric population ranges from 1.0 to 2.9% depending on the body region.(*29*) Using international reference data from the Bone Mineral Density in Childhood Study,(*30*) age-, sex-, and race-specific aBMD and BMC Z-scores were calculated and used as outcome variables.

### Demographic, Anthropometric, and Clinical Variables

Body weight (kg), height (cm) and body mass index (kg)/stature (m^2^) were obtained using standard procedures Additionally, age- and sex-specific body mass index Z-scores and categories were calculated using the international reference data for pediatric population according to the World Obesity Federation.(*31*) Somatic maturity was measured by predicting the years before or after peak height velocity (PHV) using validated algorithms for boys and girls.(*32*) Participants were categorized as pre-pubertal (< −1 year from PHV) and peri/post-pubertal (> categorized as pre-pubertal (< −1 year from PHV) and peri/post-pubertal (> −1 year 1 year from PHV).(*33*) Vitamin D status was estimated using a validated food frequency questionnaire.(*34*).(*34*) Medical records were reviewed for serum calcium levels (mg/dL), diagnosis, time from treatment completion to baseline data collection, and treatment exposure (radiotherapy, chemotherapy, and/or surgery, alone or in combination).

### Statistical Analysis

Primary and secondary outcomes were assessed using both intention-to-treat analysis, including all participants as originally randomized, and the per-protocol approach, including only adherent participants (those completing at least 95/136 sessions) with assessment at both time points (available in the supplementary online materials). Missing data were not imputed,(*35, 36*) as baseline values were considered as part of the outcome vectors, and all participants with at least one assessment were included in the analyses.(*37*) Missing data for post-intervention outcomes (both primary and secondary) were attributed to participants’ withdrawal from the study before completion and were assumed to be missing at random since there were not significant differences between dropouts and non-dropouts.

Intervention effects on primary and secondary outcomes were analyzed using a constrained longitudinal data analysis (cLDA) with linear outcomes, constraining the baseline means of the outcome measure to be the same in both groups.(*37*) The model included fixed effects for time (two levels), treatment (coded 0 for all groups at baseline and 1 or 0 at the 9-month follow-up for the exercise and control groups, respectively), and a unique participant identifier as a random effect. The effects of time and time × group interactions were also included in the model. Data are presented as within-group mean changes and between-group mean change differences with 95% confidence intervals (CI). Effect sizes were calculated according to Cohen’s d formula, interpreted according to the standard criteria: approximately 0.2, 0.5, and 0.8 standard deviations (SDs) for small effect, medium effect, and large effect sizes, respectively.(*38*) The LMMstar package was used for analysis,(*39*) and all statistical analyses were performed using R statistical software (version 4.4.0; R Foundation for Statistical Computing, Vienna, Austria). All *P*-values were obtained from two-sided tests, and *P*-values < .05 indicated statistical significance. Additionally, multiple testing corrections were applied to the primary and secondary outcomes using the false discovery rate method proposed by Benjamini and Hochberg.(*40*)

Interpretation of the intervention effect assessments was based not only on statistical significance but also on a practical benefit approach, emphasizing and reporting unadjusted values that are intuitive to human judgment and readily replicable, considering the design and methods used in this study.(*41, 42*) However, posteriori-planned subgroup analyses were performed separately by sex (boys and girls), somatic maturity (pre-pubertal and peri/post-pubertal), and low aBMD/BMC at baseline (Z-score < categorized as pre-pubertal (< −1 year from PHV) and peri/post-pubertal (> −1 year 1 and Z-score ≥ categorized as pre-pubertal (< −1 year from PHV) and peri/post-pubertal (> −1 year 1), in addition to a priori subgroup analyses by age (6–11 and 12– 18 years).

### Deviations from the original protocol

Although we originally intended to analyze primary and secondary outcomes using analysis of covariance, we were advised to use a constrained baseline longitudinal analysis via a linear mixed model as it is currently being considered the standard method and handle missing values reasonably well adopting this for primary analyses.(*37*) Nevertheless, in secondary analyses we also provide the per-protocol approach using both analysis of covariance and a constrained baseline longitudinal analysis via a linear mixed model, and the results are consistent (Supplementary Tables 4 and 5).

### List of Supplementary Materials

Table S1. CONSRT 2010 checklist.

Table S2. Descriptive characteristics of the participants that completed the study (i.e., nondropouts) and those that did not complete the study (i.e., dropouts) at completed the study at baseline.

Table S3. Distribution of cancer types of participants included in this study.

Table S4. Baseline, within-group changes (baseline to 9-month follow-up) and between-group differences (exercise vs. control) in the aBMD and BMC Z-score outcomes (per-protocol analysis).

Table S5. Baseline, within-group changes (baseline to 9-month follow-up) and between-group differences (exercise vs. control) in the aBMD and BMC Z-score outcomes (per-protocol analysis).

Fig. S1. Intention-to-treat effects by age (A), sex (B), somatic maturity (C), and low aBMD/BMC at baseline (D). Dots represent the between-groups mean difference and error bars indicate 95% confidence intervals. Data were analyzed using a constrained longitudinal data analysis (cLDA) with linear outcomes, assuming baseline means of the outcome measure were identical between groups. Age-, sex-, and race-specific aBMD and BMC Z-scores at each site are presented using international reference data from the Bone Mineral Density in Childhood Study 2. Somatic maturity groups were separately created for prepubertal (< categorized as pre-pubertal (< −1 year from PHV) and peri/post-pubertal (> −1 year 1 year from PHV) and peri/post-pubertal survivors (> categorized as pre-pubertal (< −1 year from PHV) and peri/post-pubertal (> −1 year 1 year from PHV) in accordance with Faigenbaum et al. 3. Similarly, survivors with and without low aBMD/BMC at baseline (Z-score < categorized as pre-pubertal (< −1 year from PHV) and peri/post-pubertal (> −1 year 1 and Z-score > categorized as pre-pubertal (< −1 year from PHV) and peri/post-pubertal (> −1 year 1) were stratified. Abbreviations: aBMD, areal bone mineral density; BMC, bone mineral content.

Fig. S2. Estimated means at constrained baseline and 9-month post-intervention follow-up in the aBMD and BMC Z-score outcomes. Error bars represent 95% confidence intervals. Data were analyzed using a constrained longitudinal data analysis (cLDA) with linear outcomes, assuming baseline means of the outcome measure were identical between groups (Per-protocol analysis). Abbreviations: aBMD, areal bone mineral density; BMC, bone mineral content; diff, Difference. References (only cited in Supplementary Materials)

## Supporting information

study protocol

## Data Availability

Data and materials available on request due to privacy/ethical restrictions.

## Author Contributions

Dr Andres Marmol-Perez and Dr Luis Gracia-Marco had full access to all of the data in the study and take responsibility for the integrity of the data and the accuracy of the data analysis.

Conception and design: Esther Ubago-Guisado, Vicente Martinez-Vizcaino, Francisco B Ortega, Jonatan R Ruiz, Luis Gracia-Marco

Acquisition, analysis, or interpretation of data: Andres Marmol-Perez, Esther Ubago-Guisado, Vicente Martinez-Vizcaino, Francisco B Ortega, Jonatan R Ruiz, Luis Gracia-Marco

Drafting of the manuscript: Andres Marmol-Perez.

Critical review of the manuscript for important intellectual content: Andres Marmol-Perez, Francisco B Ortega, Jonatan R Ruiz, Luis Gracia-Marco Statistical analysis: Andres Marmol-Perez, Vicente Martinez-Vizcaino, Francisco B Ortega, Jonatan R Ruiz, Luis Gracia-Marco

Administrative, technical, or material support: Andres Marmol-Perez, Esther Ubago-Guisado, Francisco B Ortega, Jonatan R Ruiz, Luis Gracia-Marco Supervision: Jonatan R Ruiz, Luis Gracia-Marco.

Final approval of manuscript: All authors.

## Data and materials availability

Data and materials available on request due to privacy/ethical restrictions.

