## Supplementary material for "Effects of a bone-strengthening exercise intervention on bone health in pediatric cancer survivors: A Randomized Controlled Trial": study protocol

### Trial Protocol and Statistical Analysis Plan

**Contact details:**


- Luis Gracia-Marco, Department of Physical and Sports Education, Faculty of Sports Science, University of Granada, Carretera de Alfacar s/n, 18071 Granada, Spain.

INDEX

1. INTRODUCTION ____ 3

1.1 Background and rationale ____ 3

1.2 Exercise and bone health ____ 4

2. AIMS ____ 4

3. METHODS/DESIGN ____ 5

3.1 Study design ____ 5

3.2 Ethical approval ____ 5

3.3 Inclusion and exclusion criteria ____ 5

3.4 Recruitment ____ 6

3.5 Participant adherence and compliance ____ 6

3.6 Intervention ____ 6

3.7 Outcomes ____ 10

4. STATISCAL ANALYSIS PLAN ____ 14

4.1 Randomization and blinding ____ 14

4.2 Sample size ____ 14

4.3 Statistical analysis ____ 15

5. SUMMARY OF CHANGES TO STUDY PROTOCOL ____ 16

### 1. INTRODUCTION

#### 1.1 Background and rationale

Owing to major advances in cancer screening and treatment over the last 30 years, cancer survival has improved dramatically. In Europe, the paediatric cancer incidence increases annually by 0.54% in children (0–14 years) and by 0.96% in adolescents (15–19 years), although it seems that the incidence in adolescents is decelerating [1]. Nevertheless, the 5-year survival rate is now at 77.9% for children and 79% for adolescents and young adults (20– 39 years) [2, 3]. Unfortunately, the treatment of paediatric cancer by means of radiation, chemotherapy and/or surgery is associated with various late effects (e.g. impaired growth, musculoskeletal sequelae, cardiopulmonary compromise and secondary malignancy) [4–6], predisposing paediatric cancer survivors to disabling conditions [4]. Furthermore, paediatric cancer treatment has been documented to have an effect on emotional well-being and quality of life, with survivors reporting anxiety, depression and post-traumatic stress [7, 8].

Paediatric cancer is a life-threatening condition that also occurs during the period of bone development and strengthening. Gonadal failure following to pelvic radiation or gonadotoxic chemotherapy and hypothalamic pituitary dysfunction by means of cranial radiation can adversely affect areal bone mineral density (aBMD), increasing osteoporosis risk later in life [9, 10]. In addition, direct radiation to bone not only causes hypovascularity but has a direct cytotoxic effect on the epiphyseal chondrocytes [11]. Observational studies have found low aBMD during and after cancer treatment to be associated with increased fracture risk (80% increase for every 1 SD reduction in lumbar spine aBMD Z-score) [12–15], which can lead to a higher risk of osteopenia and osteoporosis in adulthood and finally, disability [16]. Moreover, data from a review showed that up to 68% of paediatric cancer survivors presented moderate-to-severe aBMD deficits (Z-score $< -1$), while up to 46% had severe aBMD deficits (Z-score $< -2$) [17].

The attainment of peak bone mass during childhood and adolescence determines the aBMD later in life and therefore the onset of osteoporosis [18, 19]. This process has a strong genetic component, although lifestyle factors (i.e. physical activity and dietary habits) contribute up to 20% of the variation in peak bone mass [20]. High-intensity, weight bearing physical activity that elicit a variety of strains and include multiple rest periods is known to improve bone mass [21–24], accrual [25] and maintenance [26] as the skeleton adapts to the loads under which it is placed. Likewise, an adequate calcium and vitamin D intake in combination with physical activity is necessary to obtain beneficial gains in bone health in children and adolescents [27, 28]. In this sense, calcium and vitamin D supplementation did not add benefit to nutritional counselling for improving bone outcomes among adolescents and young adults survivors of acute lymphoblastic leukemia [29].

##### 1.2 Exercise and bone health

Exercise contributes to the development of bone mass in youths due to its association with increases in lean mass [30, 31]. Larger muscles exert greater forces on the bones, which will adapt and therefore improve their strength [32]. Furthermore, plyometric jump training is one of the best methods to improve bone health since the impacts produced against the ground will cause higher forces on the bones [33]. A recent systematic review has shown that plyometric jump training causes improvements in bone mineral content (BMC), aBMD and structural properties in children and adolescents [34]. More specifically, an 8-month jumping intervention (~ 3 min/day) improved bone mass in the proximal femur in pubertal children [35]. Mackelvie et al. [36] showed that a 7-month jumping intervention (10 min, 3 times/week) enhanced bone mass in the femoral neck and lumbar spine in pubertal girls. Additionally, Vlachopoulos et al. [37, 38] found that a 9-month jumping intervention (10 min, 3 to 4 times/week) improved bone outcomes in adolescent males participating in non-osteogenic sports and with poorer bone health.

A similar effect might be seen in survivors of paediatric cancer. A randomized controlled trial (RCT) in children with acute lymphoblastic leukemia showed that resistance exercise was unsuccessful in preventing the reduction in aBMD [39]. However, the intervention (duration, load) was not properly described. A RCT focusing on low-magnitude, high frequency mechanical stimulation seemed to improve total body aBMD in paediatric cancer survivors, while a reduction was observed in the placebo group [40]. In a recent study in children with cancer, the exercise program was not successful in improving aBMD nor other factors such as physical function or health-related quality of life [41]. This was because exercise requires of certain intensity to modify these factors, and this could not be achieved during treatment due to the child’s responses to the treatment and disease. Considering the gap in the literature, and taking into account the Exercise Guidelines for Cancer Survivors [42, 43], it is crucial to develop and implement feasible exercise program focused on improving bone health into survivorship.

#### 2. AIMS

The aim of this study is to investigate the effect of a 9-month online exercise program on bone health in paediatric cancer survivors aged 6–18 years and to follow up these outcomes 4months after the intervention to determine the extent of residual effect. We hypothesize that the intervention stimulus will be enough to improve bone health in this population. We will also examine the effect of the intervention on body composition, physical fitness, physical activity, calcium intake, vitamin D, blood samples quality of life and mental health.

#### 3. METHODS/DESIGN

##### 3.1 Study design

This protocol is reported based on Standard Protocol Items: Recommendations for Interventional Trials (SPIR IT) guidelines [44]. The iBoneFIT study is a multicenter, parallel groups RCT (1:1) designed under the equivalence basis and registered in isrctn.com (Reference: isrctn61195625, 2 April 2020). Eligible participants from two paediatric oncology units of Southern Spain will be contacted, informed, and if consenting, enrolled into the study after a meeting (T− 1) (see recruitment section). Then, randomization will be performed by an external partner who is independent of the participant recruitment and enrolment process (see randomization section). Assessments will be conducted at baseline (T0) and after nine (T1) and thirteen (T2) months in the Sport and Health University Research Institute (iMUDS, University of Granada). After finishing the study, participants in the control group will be offered the same online exercise program. A graphical description of the study design is shown in Fig. 1.

##### 3.2 Ethical approval

The study will be performed following the ethical guidelines of the Declaration of Helsinki, last modified in 2013. This study has been checked and approved by the Ethics Committee on Human Research of Regional Government of Andalusia (Reference: 4500, December 2019).

##### 3.3 Inclusion and exclusion criteria

The iBoneFIT study will include paediatric cancer survivors: 1) aged 6 to 18 years; 2) diagnosed at least 1 year earlier; 3) to have been exposed to radiotherapy and/or chemotherapy; and 4) not currently receiving treatment for cancer.

Exclusion criteria are defined as follows: 1) simultaneous participation in another study that place participants at any additional risk, discomfort or affect the results of both studies; 2) previous diagnosed anorexia nervosa/bulimia, known pregnancy and/or known alcohol and drug abuse; 3) children requiring chronic oral glucocorticoid therapy; 4) having an injury that may affect daily life activities and can be aggravated by exercise; and 5) to have a lower limb prosthesis that prevent bone assessment.

##### 3.4 Recruitment

Eligible participants will be contacted via telephone calls or information letters from the Units of Paediatric Oncology of the 'Virgen de las Nieves' (Granada) and 'Reina Sofía' (Córdoba) University Hospitals in Southern Spain. A short study information brochure will be used in routine check-ups. A meeting will be held with potential participants and parents/tutors to carefully inform about the benefits and risks of the study, and researchers will answer any question that they may have. Then, informed consents will be given, and participants will have 15 days to send it to the researchers. A hotline will be available to clarify remaining questions about the study. Those who do not react to the study invitation will be followed up via phone call at the end of these 15 days in order to check if they wish to participate. All participants will sign the informed consent before their visit to the iMUDS.

##### 3.5 Participant adherence and compliance

Participants will be allowed to withdraw at any time; nevertheless, several strategies will be used for adherence and compliance with the intervention. The minimum compliance allowed at each phase of the intervention will be 50% but the overall compliance after 9 months will have to reach 70%. A lack of compliance (<50%) without justified reasons in the first phase of the intervention will result in the participant being invited to drop out from the study. This 70% adherence rate means completing 95 sessions of 136. If a participant has not completed 70% of the intervention by the end of the 9 months but can reach 70% within two additional weeks, the exercise program will be extended for them. Compliance with the intervention will be monitored using a diary and it will be sent to the research staff on a monthly basis (Item 5). Parental involvement will be requested for this matter.

Participants and their parents are verbally motivated to participate in the intervention and to attend to all the assessments. Children who complete successfully the intervention will get a certificate of achievement. Children are the key part of this study and they deserve acknowledgements for their positive attitude and willingness (and their family) to participate in this study.

##### 3.6 Intervention

**Exercise program rationale**

The rationale of the iBoneFIT exercise program will be described following the Consensus on Exercise Reporting Template (CERT) criteria recommendations [45]. The items detailing the recommendations are shown in Table 1.

Since plyometric jump training has been shown to be effective in improving bone health and to maintain the benefits after the intervention in children and adolescents [34], jumping exercise will be the basis for the specific exercise type in iBoneFIT. Notwithstanding, the Exercise Guidelines for Cancer Survivors recommend an extended phase of resistance training before progressing to impact loading [43]. In this sense, a recent systematic review highlighted that resistance training should be incorporated at an early age and prior to plyometric training in order to establish an adequate foundation of strength for power training activities [46]. Therefore, all participants will start with a familiarization phase aimed to improve muscular fitness before implementing mechanical loading through jumps (Item 7a and 15).

Although the duration of the jumping interventions to be effective on bone outcomes in children and adolescents is unclear, the length of the exercise program will be 9 months based on results from previous studies [37, 47]. In addition, we have considered the fact that bone remodeling process requires approximately 5 months [48]. Dietary counselling on calcium and vitamin D will be provided to the participants in both control and intervention groups due to having an adequate calcium and vitamin D levels is important as both interact with physical activity to enhance bone mass (Item 9) [27, 49].

**Exercise program characteristics**

This home-based intervention will be delivered online by making use of social media (Item 4 and 12). Using popular existing social network sites may address issues of reach, engagement, and retention [50, 51]. WhatsApp (WhatsApp Inc., Mountain View, CA, USA) is a highly used app in Spain for social networking and that allows us to send text messages and other types of media (e.g. photos and videos) to the parents of participants. Although WhatsApp has been revealed as a feasible method to deliver exercise interventions, Muntaner-Mas et al. [52] have suggested that the implementation of behavior change techniques could increment the effectiveness on the outcomes assessed. Thus, five behavior change techniques (i.e. action planning and goal setting, providing instructions and demonstrations of how to perform the behavior, self-monitoring of behavior, providing feedback on performance and information about health consequences) and a gamification design (i.e. points and rankings) will be included to improve the interest and incentive of this non-game program (Table 2) (Item 6). These motivational approaches were chosen because of their known effect on physical fitness

[52], physical activity [53] and satisfaction [54]. Moreover, parents will be told to encourage their children to perform the exercise program in order to increase motivation.

A personal trainer with a BSc degree in Sport Sciences will develop all the sessions of this program (Item 2, 14a and 14b). The personal trainer will record 18 exercise sessions and they will be uploaded in a private channel of the YouTube website. Each of them will be repeated over a 2 weeks period. The YouTube platform has been reported to be an educational tool for health-care conditions among people coping with illness [55]. Every new session for the following 2 weeks will be shared through the WhatsApp group every 2 weeks. Finally, participants will perform the exercise program individually or accompanied (i.e. with parents or friends) according to their preferences (Item 3) [56]. They will be required to record videos and send through the WhatsApp group in order to supervise the execution of the jumping exercises by the personal trainer. The exercise program will be performed on a hard surface (Item 1) [57], and participants will be asked to report any pain or injuries at each stage of the intervention (Item 11).

**Frequency and volume**

Following the updated physical activity guidelines, children and adolescents should include bone-strengthening exercises as part of the daily physical activity on at least 3 days per week. Participants in the iBoneFIT study will perform the exercise program three to 4 days per week (preferably on Mondays, Wednesdays and Fridays; or Mondays, Tuesdays, Thursdays and Fridays). If one training session is missed, the participant will be able to do it on a different day of the week, provided a minimum of 24 h of rest.

The total volume will be 7296 squat/jumps (2000 squats +5296 jumps). The doses will be composed of 136 sessions (10–20 min/session) over 36 weeks. A full description of the training volume and its progression is shown in Table 3. In a recent 9-month RCT based on jumping activities with similar dosage we reached 70% of compliance (6216 jumps), and this was enough to improve bone outcomes in non-weight-bearing sport athletes [37]. Thus, the proposed volume of 7296 squat/ jumps is likely to elicit the same effect in paediatric cancer survivors.

**Periodization**

Although Peitz et al. [58] did not find differences between no, linear and undulating periodisations in youth, iBoneFIT will implement a linear model based on the fact that variation in volume and/or impact loading within the program phases may stimulate greater bone adaptions and reduce boredom and risk of overtraining [43]. The exercise program will be divided in three phases of different durations and impact loadings (i.e. height reached in the different jumps). Each phase will be composed of levels with progressive increase in volume (i.e. repetitions, sets per day and sessions per week) as shown in Table 3 (Item 7b).

The phase 1 corresponds to the first 8 weeks of the exercise program. Participants will perform body mass-based squats and the volume will increase progressively by modifying the number of repetitions and sets per day. Paediatric cancer survivors may present reduced aBMD and muscular fitness [59], therefore jumping exercise prescription may not be safe. In this sense, body mass-based squat was chosen in this phase following previous studies that observed positive effects on muscular fitness after an 8-week intervention [60, 61].

The phase 2 will last 12 weeks and participants will perform squat jumps. In this phase, the volume will increase progressively by modifying the number of repetitions, sets per day and sessions per week. Squat jump has been chosen as intermediary exercise before the use of countermovement jump since the jump height reached is lower and hence, ground reaction forces produced at the landing are lower [62]. Furthermore, squat jump training reduces the degree of muscle slack on the push-off phase [63] which could supply a better execution of the countermovement jump afterwards.

The phase 3 will be the longest phase of the exercise program with 16 weeks. Participants will perform countermovement jumps and the volume of this phase will be increased progressively by modifying the number of repetitions, sets per day and sessions per week. Countermovement jump will be chosen in this phase since it produces a huge force application (~400 times body mass / second) and ground reaction forces (~5 times body mass) in youth [64, 65]. Countermovement jump has been previously reported to be valid and reliable in children [66].

**Session structure**

The structure of the exercise sessions will be: 1) warm up; 2) squat/jumps training; 3) cool down. Briefly, the warm ups will be based on RAMP methodology (i.e. raise, activate, mobilize and potentiate) in order to maximize middle-term performance of the main exercises (i.e. squat/jumps exercises) [67]. Eight exercises focused on the brace, squat, lunge or jump patterns will be included in this part of the session. Squat/jumps training will comprise body mass-based squats, squat jumps and countermovement jumps in phase 1, phase 2 and phase 3, respectively. Finally, participants will perform a cool down including static stretching and relaxing exercises (Item 8).

**Control group**

Participants randomly allocated to the CG will receive information on the recommendations of calcium and vitamin D [68]. Educational leaflets and infographics based on the current recommendations [68] will be delivered to the participants at the beginning of the study (Item 10). After finishing the study, they will be offered the same online exercise program.

#### 3.7 Outcomes

The primary outcome of our study is bone health. The secondary outcomes include anthropometric measurements, body composition, physical fitness components, free-living physical activity, blood samples, calcium and vitamin D intake, health-related quality of life and mental health. Assessments will be conducted at baseline, repeated at post-test (i.e. after 2 weeks of intervention or control condition at most) and follow up (i.e. after 4 months of intervention or control condition). Participants will be assessed for the post-test and follow up following the order through which they will be tested at baseline, to avoid cofounding by time between baseline and the other assessments.

Data obtained on the assessments will be recorded on a paper print-out and entered into an Excel file for future statistical analysis. Questionnaires will be filled using Google Forms which allows us to record the data without hand-written management. In compliance with the Personal Information Protection Act, the names of all participants will not be disclosed, and an identifier number will be used to identify each participant. All participants will be informed that the clinical data obtained in the trial will be stored in a computer and will be handled with confidentiality.

**Primary outcome: bone health**

**Dual-energy X-ray Absorptiometry (DXA)**

A DXA (Hologic Series Discovery QDR, Bedford, MA, USA) will be used throughout the study to obtain BMC (g) and aBMD (g/cm2) for the hip, lumbar spine and total body less head. Furthermore, lean soft tissue mass (g), fat mass (kg) and body fat percentage (%) for the whole body will be obtained from total body scans. APEX software (version 4.0.2) will be used to analyze the scans following the recommendations for children and adolescents [69]. Equipment calibration, participant setting and scan analyses will be performed by the same researcher. DXA uses a minimal radiation (i.e. spending a day outside in the sunshine) and the effective dose for the scans in children has been set in 3–6 μSv [70].

Hip Structural Analysis (HSA) HSA is a DXA-based software that analyses hip scans to estimate bone geometric properties of the proximal femur. This software analyses structural characteristics through the distribution of bone mineral mass in a line of pixels across the bone axis [71]. These geometric estimates in the proximal femur will be derived from: 1) the cross-sectional area ($mm^2$); 2) section modulus ($mm^3$); and 3) the cross-sectional moment of inertia ($mm^4$). For these variables, the short-term coefficient of variation has been reported to be between 2.4 and 10.1% [72].

Trabecular Bone Score (TBS) TBS is a DXA-based software (iNsight version 3.0, Medimaps, Pessac, France) that indirectly assesses the state of trabecular microarchitecture in the lumbar spine. Based on experimental variograms of the projected DXA image, TBS evaluates the heterogeneity of the grey-levels pixels of the aBMD and higher heterogeneity implies worse trabecular connectivity [73]. Low values reported in this parameter have been associated with a higher fracture risk, and therefore it is considered an index of bone quality [74]. The short-term coefficient of variation for TBS has been reported to be between 1.7 and 2.1% for spine aBMD in 92 individuals with repeated spine DXA scans performed within 28 days [75]. 3D-DXA Modelling 3D-SHAPER is a DXA-based software (version 2.2, Galgo Medical, Barcelona, Spain) that derives 3D analyses from the hip DXA scans. Details of the model algorithm are published elsewhere [76]. Briefly, this software uses a 3D statistical shape and density of the proximal femur built from a database of quantitative computed tomography (QCT) scans of Caucasian population [76]. The 3D-SHAPER will assess bone parameters such as the cortex, the femoral shape and the trabecular macrostructure [77]. The cortex is segmented by fitting a mathematical function of the cortical thickness (mm), cortical volumetric BMD (cortical vBMD, mg/cm3), the location of the cortex, the density of surrounding tissues and the imaging blur to the density profile computed along the normal vector at each node of the proximal femur surface mesh [77]. In addition, the cortical surface BMD (cortical sBMD, mg/cm2) is computed at each vertex of the femoral surface mesh, as the multiplication of the cortical thickness (cm) by the cortical vBMD along its thickness [78]. Any increase in either cortical thickness or cortical vBMD will ensure an increase in cortical sBMD. Nevertheless, if cortical thickness and cortical vBMD vary in opposite ways, cortical sBMD will remain unchanged. All measurements will be computed over the total femur (i.e. the shaft, the intertrochanteric and the union of the neck) according to the trabecular, cortical and integral compartments. Correlation coefficients between BMD computed by 3D-SHAPER and QCT of the total femur have been reported to be 0.86–0.95, whereas the correlation coefficients of BMD computed by 3D-SHAPER with BMD computed by QCT have been reported to be 0.91

[76]. The short-term coefficients of variations of aBMD measurements have been reported to be 1.5, 4.5, 1.7 and 1.5% for cortical thickness, trabecular vBMD, cortical vBMD and cortical sBMD, respectively [78].

**Secondary outcomes**

**Anthropometric measurement, body composition and somatic maturation**

Body mass (kg) will be measured with an electronic scale (SECA 861, Hamburg, Germany) with an accuracy of 100 g. Height (cm) will be measured by using a precision stadiometer (SECA 225, Hamburg, Germany) to the nearest 0.1 cm. Body mass index (BMI) will be calculated as body mass (kg)/height (m2), and the participants will be classified into BMI categories according to sex- and age-specific cut offs [79].

In addition to DXA measurements, a bioimpedance scale (Tanita BC-418 MA; Amsterdam, The Netherlands; range: 2–200 kg; precision: 0.1 kg; body fat percentage increments: 0.1%) will estimate the percentage of body fat of the participants. The assessment will be carried out in fasting state according to the manufacturer's instructions. Despite the measured error, bioelectrical impedance analysis will be used to assess body fat as it is considered a practical method in addition to DXA [80]. Somatic maturation will be assessed using the prediction of years from peak height velocity using validated algorithms for children [81].

**Physical fitness**

The ALPHA fitness test battery will be used to assess physical fitness. These field-based fitness tests have been shown to be valid, reliable and related to health in children and adolescents [82]. In brief, cardiorespiratory fitness will be assessed with the 20m shuttle run test; muscular fitness will be assessed with the handgrip strength and standing long jump tests; and speed agility will be assessed with the 4 × 10 m shuttle run test. All tests will be performed twice, and the best score will be retained, except 20 m shuttle run test.

Perceived physical fitness will be assessed by the International Fitness Scale (IFIS). The IFIS is a short, simple and self-administered scale that has been validated in children and adolescents [83, 84]. This 5-item scale asks the participants about their physical fitness comparing with their colleagues.

**Physical activity and sedentarism**

Physical activity and sedentary behaviors will be objectively assessed at the baseline, post-intervention and follow-up measurements. Participants will wear a tri-axial accelerometer

(ActiGraph GT3X, Pensacola, FL, USA) attached to the non-dominant wrist over seven consecutive days (24 h/day) and they will remove it only for water-based activities (e.g. bathing or swimming). They will also have a diary in order to record the time when they go to bed, wake up and remove the device. Correlation coefficient between accelerometer measured metabolic energy equivalents and indirect calorimetry has been reported to be 0.65 [85], whilst correlation coefficient of accelerometer impact loading and ground reaction forces by force platforms has been reported to be 0.74 [86].

In addition, information on self-reported physical activity and sedentary behaviors will be obtained by the cross-translated and adapted version of the Youth Activity Profile (YAP) questionnaire (available at: http://profith.ugr.es/ yap?lang=en). The YAP questionnaire was developed at the Iowa State University and validated in children [87]. This self-administered 7-day recall questionnaire collects data from items regarding physical activity in the school setting, physical activity out of the school setting, activity immediately after school, activity during the evening and activity during each weekend day. Moreover, the bone-specific physical activity questionnaire (BPAQ) will be used to assess the influence of historical physical activity (i.e. activities in which you have ever participated, and activities practiced in the last 12months) on skeletal health. It has been reported that BPAQ is a valid instrument to account for the effects of previous physical activities on the skeleton [65].

**Calcium intake and vitamin D status**

To correctly interpret bone health of the participants, an assessment of dietary intake of calcium will completed at the baseline, post-intervention and follow-up measurements. A validated food-frequency questionnaire will be used to estimate calcium intake [88]. In addition to plasma 25-hydroxyvitamin D levels obtained from blood analyses (see blood samples section), a vita- min D questionnaire to assess the status of this prohormone will be implemented [89].

**Blood samples**

Fasting blood samples will be collected by venepuncture between 8:00 and 10:00 after an overnight fast. The methodology for shipment, preparation and collection of the blood samples was standardized among all participating hospitals. A set of parameters obtained from hematological and biochemical analyses will be available from the hospitals as part of the follow-up protocols.

**Health-related quality of life and mental health**

The Paediatric Quality of Life Inventory (PedsQLTM 4.0 Generic Core Scales) will be used to assess quality of life. PedsQLTM is validated in paediatric cancer survivors and has been successfully used [90]. This 23-item scale assesses quality of life considering five domains of health (i.e. physical functioning, emotional functioning, psychosocial functioning, social functioning and school functioning). Results from our participants in all domains of PedsQLTM will be compared to published normative data [91].

Childhood anxiety will be assessed with the State-Trait Anxiety Inventory for Children (STAIC-T). This inventory has been extensively validated in Spanish children [92]. Depression will be measured with the Children Depression Inventory (CDI), which consists of 27 items that assesses 5 domains (interpersonal problems, ineffectiveness, negative mood, anhedonia and negative self-esteem) [93]. Rosenberg Self-Esteem scale will be used to assess self-esteem and has been validated with children and adolescents [94]. We will use the Positive Affect Schedule for children (PANAS-C) in order to measure both positive and negative affect [95]. The original PANAS-C reported appropriate values of internal consistency (0.86 for the positive affect and 0.82 for the negative affect). Happiness will be assessed by the Subjective Happiness Scale (SHS) whose Spanish version has shown appropriate test-retest reliability, internal consistency and convergent validity [96]. Dispositional optimism will be assessed with the Life Orientation Test-Revised (LOT-R) [97]. LOT-R is an instrument with good internal consistency (0.71 for the total score and of 0.64 and 0.77 for the optimism and pessimism, respectively) [98].

#### 4. STATISCAL ANALYSIS PLAN

##### 4.1 Randomization and blinding

Randomization to an intervention group (online exercise program, IG) or control group (no treatment, CG) will be performed by an external partner (V.M-V) who is independent of the participant recruitment and enrolment process, stratified by age and sex. Each participant will be provided a uniform (0, 1) random number using SAS software, version 9.1 (SAS Institute Inc), within their respective age and sex group. Assignments will be blinded to the assessors until all tests are completed. For feasibility reasons, the study will probably be conducted in two waves of 58 children at most.

##### 4.2 Sample size

We have used femoral neck aBMD as the outcome to calculate the sample size, since it is a key variable in the diagnosis of osteoporosis. Since the study will include children and adolescents (6–18years), the sample size has been calculated taking into account that sub-group analysis by age groups (6 to 11 years and 12 to 18 years) may be required. Based on an expected effect size of 0.25 for the change in femoral neck aBMD, an $\alpha$ level of 0.05 and a power of 80%, a minimum of 116 participants will be required (IG = 58 and CG = 58). This includes a 20% extra for occasional losses and refusals and 10% for multivariable analyses. Calculations have been obtained using G*Power (v.3.1.9.2) with analysis of variance: repeated measures (within-between interactions) for 2 groups (between factors) and 2 time points (pre, post, within factors). A correlation between measures of 0.7 has been assumed, which is achievable when measuring bone outcomes [99].

**4.3 Statistical analysis**

All variables will be checked for normality using both statistical and graphical methods. Results will be presented as frequencies and proportions with 95% confidence intervals for categorical variables and mean (standard deviation) or median (range) for continuous variables. A descriptive analysis of the participants characteristics will be performed as soon as the baseline assessments are completed. Intervention effects on bone outcomes (age and sex-adjusted Z-scores) will be analyzed using constrained baseline longitudinal analysis via a linear mixed model. As baseline values will be considered as part of the outcome vectors, all survivors with at least one evaluation (baseline or follow-up) will be included in the analyses. The model will include fixed effects for time (two levels), treatment (coded 0 for all groups at baseline and coded 0 or 1 at follow-up for the exercise and control groups, respectively), as well as the unique survivor identifier as a random effect. Data will be presented as within-group mean changes and differences between-group mean changes with 95% confidence intervals unless mentioned. The LMMstar package will be used to construct the linear mixed models for the analysis. Statistical analyses will be performed using the statistical software R version 4.0.3 (R Foundation for Statistical Computing).

#### 5. SUMMARY OF CHANGES TO STUDY PROTOCOL

| Date | Amendment |
|---|---|
| 01/12/2019 | Approval by the Ethics Committee on Human Research of Regional Government of Andalusia (Reference: 4500). |
| 02/04/2020 | First posted at isrctn.com (Reference: isrctn61195625). |
| 01/10/2020 | Presentations of press releases to emphasize the importance of calcium and vitamin D for bone health were not given at the beginning of each phase.<br><br>This modification was made to place the same emphasis on calcium and vitamin D importance in both groups at baseline. |
| 05/02/2024 | Intervention effects on primary and secondary end points were analyzed using a constrained baseline longitudinal analysis via a linear mixed model.<br><br>Missing data for post-intervention end points were attributed to participants' withdrawal from the study before completion and were assumed to be missing at random since there were not significant differences between dropouts and non-dropouts. Therefore, we were advised to use a constrained baseline longitudinal analysis via a linear mixed model as it is currently being considered the standard method and handle missing values reasonably well [100]. Nevertheless, results were consistent when using constrained baseline longitudinal analysis via a linear mixed model and analysis of covariance (data not shown) in the per-protocol approach. |
