## supplementary online materials for "Effects of a bone-strengthening exercise intervention on bone health in pediatric cancer survivors: A Randomized Controlled Trial"

***Supplementary Information***

**Table S1.** CONSRT 2010 checklist.

| Section/Topic | Item No | Checklist item | Reported on page No |
|---|---|---|---|
| **Title and abstract** | | | |
| | 1a | Identification as a randomised trial in the title | 1 |
| | 1b | Structured summary of trial design, methods, results, and conclusions (for specific guidance see CONSORT for abstracts) | 3-4 |
| **Introduction** | | | |
| Background and objectives | 2a | Scientific background and explanation of rationale | 5 |
| | 2b | Specific objectives or hypotheses | 5-6 |
| **Methods** | | | |
| Trial design | 3a | Description of trial design (such as parallel, factorial) including allocation ratio | 14 |
| | 3b | Important changes to methods after trial commencement (such as eligibility criteria), with reasons | 21 |
| Participants | 4a | Eligibility criteria for participants | 14-15 |
| | 4b | Settings and locations where the data were collected | 14-15 |
| Interventions | 5 | The interventions for each group with sufficient details to allow replication, including how and when they were actually administered | 16-17 |
| Outcomes | 6a | Completely defined pre-specified primary and secondary outcome measures, including how and when they were assessed | 17-19 |
| | 6b | Any changes to trial outcomes after the trial commenced, with reasons | - |
| Sample size | 7a | How sample size was determined | 14-15 |
| | 7b | When applicable, explanation of any interim analyses and stopping guidelines | - |
| Randomisation: | | | |
| Sequence generation | 8a | Method used to generate the random allocation sequence | 15 |
| | 8b | Type of randomisation; details of any restriction (such as blocking and block size) | 15 |
| Allocation concealment mechanism | 9 | Mechanism used to implement the random allocation sequence (such as sequentially numbered containers), describing any steps taken to conceal the sequence until interventions were assigned | 15 |
| Implementation | 10 | Who generated the random allocation sequence, who enrolled participants, and who assigned participants to interventions | 15 |

| | | | |
|---|---|---|---|
| Blinding | 11a | If done, who was blinded after assignment to interventions (for example, participants, care providers, those assessing outcomes) and how | 15 |
| | 11b | If relevant, description of the similarity of interventions | - |
| Statistical methods | 12a | Statistical methods used to compare groups for primary and secondary outcomes | 19-21 |
| | 12b | Methods for additional analyses, such as subgroup analyses and adjusted analyses | 19-21 |
| **Results** | | | |
| Participant flow (a diagram is strongly recommended) | 13a | For each group, the numbers of participants who were randomly assigned, received intended treatment, and were analysed for the primary outcome | 6-7 |
| | 13b | For each group, losses and exclusions after randomisation, together with reasons | 6-7 |
| Recruitment | 14a | Dates defining the periods of recruitment and follow-up | 14 |
| | 14b | Why the trial ended or was stopped | - |
| Baseline data | 15 | A table showing baseline demographic and clinical characteristics for each group | 6-7 |
| Numbers analysed | 16 | For each group, number of participants (denominator) included in each analysis and whether the analysis was by original assigned groups | 6-7 |
| Outcomes and estimation | 17a | For each primary and secondary outcome, results for each group, and the estimated effect size and its precision (such as 95% confidence interval) | 7-8 |
| | 17b | For binary outcomes, presentation of both absolute and relative effect sizes is recommended | - |
| Ancillary analyses | 18 | Results of any other analyses performed, including subgroup analyses and adjusted analyses, distinguishing pre-specified from exploratory | 7-8 |
| Harms | 19 | All important harms or unintended effects in each group (for specific guidance see CONSORT for harms) | 8 |
| **Discussion** | | | |
| Limitations | 20 | Trial limitations, addressing sources of potential bias, imprecision, and, if relevant, multiplicity of analyses | 11-12 |
| Generalisability | 21 | Generalisability (external validity, applicability) of the trial findings | 12-13 |
| Interpretation | 22 | Interpretation consistent with results, balancing benefits and harms, and considering other relevant evidence | 9-13 |
| **Other information** | | | |
| Registration | 23 | Registration number and name of trial registry | 14 |
| Protocol | 24 | Where the full trial protocol can be accessed, if available | 14 |
| Funding | 25 | Sources of funding and other support (such as supply of drugs), role of funders | 33 |

**Table S2.** Descriptive characteristics of the participants that completed the study (i.e., nondropouts) and those that did not complete the study (i.e., dropouts) at completed the study at baseline.

| | Total | N | Nondropouts | N | Dropouts | N | *P* |
|---|---|---|---|---|---|---|---|
| Sex (female, %) | 42.2 | 116 | 44.4 | 99 | 29.4 | 17 | .856 |
| Age (years) | 12.1 (3.3) | 116 | 12.12 (3.42) | 99 | 11.94 (2.86) | 17 | .838 |
| Body mass (kg) | 46.6 (18.0) | 116 | 46.38 (17.76) | 99 | 47.89 (20.18) | 17 | .750 |
| Stature (cm) | 147.5 (17.1) | 116 | 147.37 (17.00) | 99 | 148.01 (18.01) | 17 | .888 |
| Body mass index Z-score | 0.9 (1.1) | 116 | 0.87 (1.11) | 99 | 1.01 (1.27) | 17 | .623 |
| Body mass index (categories, %) | | | | | | | |
| Underweight | 3.5 | 4 | 3.0 | 3 | 5.9 | 1 | .281 |
| Normoweight | 61.2 | 71 | 64.7 | 64 | 41.2 | 7 | |
| Overweight | 20.7 | 24 | 18.2 | 18 | 35.3 | 6 | |
| Obesity | 14.6 | 17 | 14.1 | 14 | 17.6 | 3 | |
| Years from peak height velocity | -0.8 (2.7) | 116 | -0.70 (2.78) | 99 | -1.07 (2.42) | 17 | .613 |
| Vitamin D deficiency (yes, %) | 53.2 | 111 | 52.6 | 97 | 57.4 | 14 | .590 |
| Calcium intake (mg/dL) | 9.8 (0.5) | 103 | 9.85 (0.50) | 87 | 9.73 (0.62) | 16 | .394 |
| Time from cancer treatment completion (years) | 5.0 (3.8) | 113 | 5.22 (3.85) | 97 | 3.86 (3.25) | 16 | .187 |
| Radiotherapy exposure (yes, %) | 27.6 | 116 | 28.3 | 99 | 23.5 | 17 | **.014** |
| aBMD Z-score | | | | | | | |
| Femoral neck | -0.2 (1.4) | 115 | -0.12 (1.36) | 98 | -0.68 (1.54) | 17 | .128 |
| Total hip | 0.1 (1.3) | 115 | 0.15 (1.27) | 98 | 0.06 (1.45) | 17 | .795 |
| Total body (less head) | -0.2 (1.4) | 116 | -0.21 (1.30) | 99 | -0.07 (1.82) | 17 | .691 |
| Lumbar spine | -0.1 (1.3) | 116 | -0.10 (1.37) | 99 | 0.08 (1.18) | 17 | .613 |
| BMC Z-score | | | | | | | |
| Femoral neck | -1.3 (1.5) | 115 | -1.30 (1.47) | 98 | -1.10 (1.73) | 17 | .617 |
| Total hip | 0.4 (1.4) | 115 | 0.35 (1.39) | 98 | 0.71 (1.58) | 17 | .333 |
| Total body (less head) | -0.5 (1.3) | 116 | -0.54 (1.27) | 99 | -0.18 (1.44) | 17 | .292 |
| Lumbar spine | -0.5 (1.3) | 116 | -0.46 (1.24) | 99 | -0.36 (1.48) | 17 | .768 |

Data are presented as mean (standard deviation) or as frequencies (percentages), as indicated. Baseline differences between nondropouts and dropouts were determined by one-way analysis of variance (ANOVA) and chi- squared tests for continuous and categorical variables, respectively. Statistically significant values at $P < .05$ are shown in bold.

Age- and sex-specific BMI Z-score and categories are presented using international reference data for pediatric population according to the World Obesity Federation *(1)*. Age-, sex-, and race-specific aBMD and BMC Z-scores at each site are also presented using international reference data from the Bone Mineral Density in Childhood Study *(2)*. Abbreviations: aBMD, areal bone mineral density; BMC, bone mineral content.

**Table S3.** Distribution of cancer types of participants included in this study.

| | Total | N | Exercise | N | Control | N |
|---|---|---|---|---|---|---|
| Type of cancer (%) | | | | | | |
| Acute lymphoblastic leukemia | 38.8 | 45 | 39.7 | 23 | 37.9 | 22 |
| Lymphoma | 12.1 | 14 | 8.6 | 5 | 15.5 | 9 |
| Central nervous system | 9.5 | 11 | 10.3 | 6 | 8.6 | 5 |
| Renal tumor | 7.8 | 9 | 6.9 | 4 | 8.6 | 5 |
| Neuroblastoma | 6.9 | 8 | 8.6 | 5 | 5.2 | 3 |
| Malignant bone tumor | 6.9 | 8 | 6.9 | 4 | 6.9 | 4 |
| Histiocytosis | 5.2 | 6 | 6.9 | 4 | 3.4 | 2 |
| Soft tissue and other extraosseous sarcomas | 4.3 | 5 | 5.2 | 3 | 3.4 | 2 |
| Retinoblastoma | 3.4 | 4 | 3.4 | 2 | 3.4 | 2 |
| Hepatic tumor | 2.6 | 3 | 1.7 | 1 | 3.4 | 2 |
| Other malignant epithelial neoplasms | 1.7 | 2 | 1.7 | 1 | 3.4 | 2 |
| Unknown | 0.9 | 1 | 0.0 | 0 | 0.0 | 0 |

Data are presented as percentages and frequencies.

**Table S4.** Baseline, within-group changes (baseline to 9-month follow-up) and between-group differences (exercise vs. control) in the aBMD and BMC Z-score outcomes (per-protocol analysis).

| | Exercise | | Control | | Between-group Differences | | |
|---|---|---|---|---|---|---|---|
| | Mean (95% CI) | Change %$^{\Psi}$ | Mean (95% CI) | Change %$^{\Psi}$ | MD (95% CI) | Effect size | *P*[†] |
| Femoral neck aBMD Z-score | | | | | | | |
| Baseline | -0.05 (-0.34 to 0.24) | NA | -0.05 (-0.34 to 0.24) | NA | NA | NA | NA |
| Change at 9-month | 0.08 (-0.11 to 0.28) | 2.70 | 0.14 (-0.02 to 0.30) | 4.05 | -0.06 (-0.27 to 0.15) | -0.12 | .598 |
| Total hip aBMD Z-score | | | | | | | |
| Baseline | 0.23 (-0.04 to 0.50) | NA | 0.23 (-0.04 to 0.50) | NA | NA | NA | NA |
| Change at 9-month | 0.20 (-0.08 to 0.47) | 4.82 | -0.04 (-0.27 to 0.18) | -2.41 | 0.24 (-0.06 to 0.53) | 0.35 | .111 |
| Total body aBMD Z-score | | | | | | | |
| Baseline | -0.19 (-0.47 to 0.10) | NA | -0.19 (-0.47 to 0.10) | NA | NA | NA | NA |
| Change at 9-month | 0.05 (-0.18 to 0.27) | 3.80 | -0.06 (-0.24 to 0.13) | -3.80 | 0.10 (-0.14 to 0.35) | 0.19 | .409 |
| Lumbar spine aBMD Z-score | | | | | | | |
| Baseline | -0.05 (-0.35 to 0.25) | NA | -0.05 (-0.35 to 0.25) | NA | NA | NA | NA |
| Change at 9-month | -0.05 (-0.20 to 0.10) | -2.70 | -0.02 (-0.14 to 0.10) | -2.70 | -0.03 (-0.19 to 0.13) | -0.08 | .704 |
| Femoral neck BMC Z-score | | | | | | | |
| Baseline | -1.25 (-1.57 to -0.93) | NA | -1.25 (-1.57 to -0.93) | NA | NA | NA | NA |
| Change at 9-month | 0.47 (0.04 to 0.91) | 13.09 | 0.00 (-0.36 to 0.36) | 7.27 | 0.48 (0.02 to 0.94) | 0.43 | **.043** |
| Total hip BMC Z-score | | | | | | | |
| Baseline | 0.41 ( 0.11 to 0.71) | NA | 0.41 ( 0.11 to 0.71) | NA | NA | NA | NA |
| Change at 9-month | 0.39 (-0.06 to 0.84) | 13.17 | -0.06 (-0.44 to 0.31) | -8.70 | 0.45 (-0.01 to 0.92) | 0.39 | .057 |
| Total body BMC Z-score | | | | | | | |
| Baseline | -0.50 (-0.78 to -0.23) | NA | -0.50 (-0.78 to -0.23) | NA | NA | NA | NA |
| Change at 9-month | 0.17 (-0.03 to 0.37) | 8.59 | -0.04 (-0.20 to 0.13) | -7.05 | 0.21 (-0.01 to 0.42) | 0.42 | .058 |
| Lumbar spine BMC Z-score | | | | | | | |
| Baseline | -0.45 (-0.72 to -0.18) | NA | -0.45 (-0.72 to -0.18) | NA | NA | NA | NA |
| Change at 9-month | 0.17 (-0.02 to 0.37) | 9.02 | -0.04 (-0.20 to 0.12) | -7.00 | 0.21 (0.00 to 0.42) | 0.42 | .050 |

Results are presented as mean change from baseline for each group and as mean difference between groups change. Data were analyzed using a constrained longitudinal data analysis (cLDA) with linear outcomes, assuming baseline means of the outcome measure were identical between groups. Age-, sex-, and race-specific aBMD and BMC Z-scores at each site are presented using international reference data from the Bone Mineral Density in Childhood Study *(2)*. Effect sizes were calculated following Cohen's d formula. Abbreviations: aBMD, areal bone mineral density; BMC, bone mineral content; CI, confidence intervals; MD, mean difference; NA, not applicable.

$^{\Psi}$ Change percentage (follow-up minus baseline values/baseline values * 100) was calculated from raw values (g/cm$^2$ or g).

[†] P-values showed between-group differences on the aBMD and BMC Z-score outcomes.

**Table S5.** Baseline, within-group changes (baseline to 9-month follow-up) and between-group differences (exercise vs. control) in the aBMD and BMC Z-score outcomes (per-protocol analysis).

| | Exercise | | Control | | Between-group Differences | | |
|---|---|---|---|---|---|---|---|
| | Mean (95% CI) | Change %$^{\Psi}$ | Mean (95% CI) | Change %$^{\Psi}$ | MD (95% CI) | Effect size | $P^{\dagger}$ |
| Femoral neck aBMD Z-score | | | | | | | |
| Baseline | 0.24 (-0.29 to 0.78) | NA | -0.25 (-0.58 to 0.08) | NA | NA | NA | NA |
| Change at 9-month | 0.08 (-0.08 to 0.24) | 2.70 | 0.14 (0.00 to 0.27) | 4.05 | -0.06 (-0.27 to 0.16) | -0.13 | .601 |
| Total hip aBMD Z-score | | | | | | | |
| Baseline | 0.47 (0.01 to 0.93) | NA | 0.06 (-0.27 to 0.39) | NA | NA | NA | NA |
| Change at 9-month | 0.20 (-0.03 to 0.42) | 4.82 | -0.04 (-0.23 to 0.14) | -2.41 | 0.24 (-0.06 to 0.54) | 0.35 | .113 |
| Total body aBMD Z-score | | | | | | | |
| Baseline | 0.12 (-0.39 to 0.64) | NA | -0.39 (-0.72 to -0.07) | NA | NA | NA | NA |
| Change at 9-month | 0.05 (-0.14 to 0.23) | 3.80 | -0.06 (-0.21 to 0.10) | -3.80 | 0.10 (-0.14 to 0.35) | 0.19 | .412 |
| Lumbar spine aBMD Z-score | | | | | | | |
| Baseline | 0.29 (-0.16 to 0.75) | NA | -0.28 (-0.67 to 0.11) | NA | NA | NA | NA |
| Change at 9-month | -0.05 (-0.18 to 0.08) | -2.70 | -0.02 (-0.12 to 0.08) | -2.70 | -0.03 (-0.19 to 0.13) | -0.08 | .705 |
| Femoral neck BMC Z-score | | | | | | | |
| Baseline | -1.14 (-1.68 to -0.60) | NA | -1.32 (-1.72 to -0.92) | NA | NA | NA | NA |
| Change at 9-month | 0.47 (0.11 to 0.83) | 13.09 | -0.01 (-0.30 to 0.28) | 7.27 | 0.48 (0.01 to 0.94) | 0.45 | **.044** |
| Total hip BMC Z-score | | | | | | | |
| Baseline | 0.61 (0.13 to 1.09) | NA | 0.28 (-0.11 to 0.67) | NA | NA | NA | NA |
| Change at 9-month | 0.38 (0.02 to 0.74) | 13.17 | -0.07 (-0.37 to 0.22) | -8.70 | 0.45 (-0.02 to 0.92) | 0.42 | .059 |
| Total body BMC Z-score | | | | | | | |
| Baseline | -0.22 (-0.64 to -0.20) | NA | -0.69 (-1.05 to -0.33) | NA | NA | NA | NA |
| Change at 9-month | 0.17 (0.03 to 0.34) | 8.59 | -0.04 (-0.17 to 0.10) | -7.05 | 0.21 (-0.01 to 0.42) | 0.42 | .060 |
| Lumbar spine BMC Z-score | | | | | | | |
| Baseline | -0.20 (-0.58 to 0.19) | NA | -0.62 (-1.00 to -0.25) | NA | NA | NA | NA |
| Change at 9-month | 0.17 (0.10 to 0.33) | 9.02 | -0.04 (-0.17 to 0.09) | -7.00 | 0.21 (-0.01 to 0.42) | 0.43 | .051 |

Results are presented as mean change from baseline for each group and as mean difference between groups change. Data were analyzed using analysis of covariance, with bone outcomes as dependent variables in separate models, group (exercise vs control) as a fixed factor, and the baseline of the study outcome as a covariate. Age-, sex-, and race-specific aBMD and BMC Z-scores at each site are presented using international reference data from the Bone Mineral Density in Childhood Study *(2)*. Effect sizes were calculated following Cohen's d formula. Abbreviations: aBMD, areal bone mineral density; BMC, bone mineral content; CI, confidence intervals; MD, mean difference; NA, not applicable.

$^{\Psi}$ Change percentage (follow-up minus baseline values/baseline values * 100) was calculated from raw values (g/cm$^2$ or g).

$^{\dagger}$ P-values showed between-group differences on the aBMD and BMC Z-score outcomes.

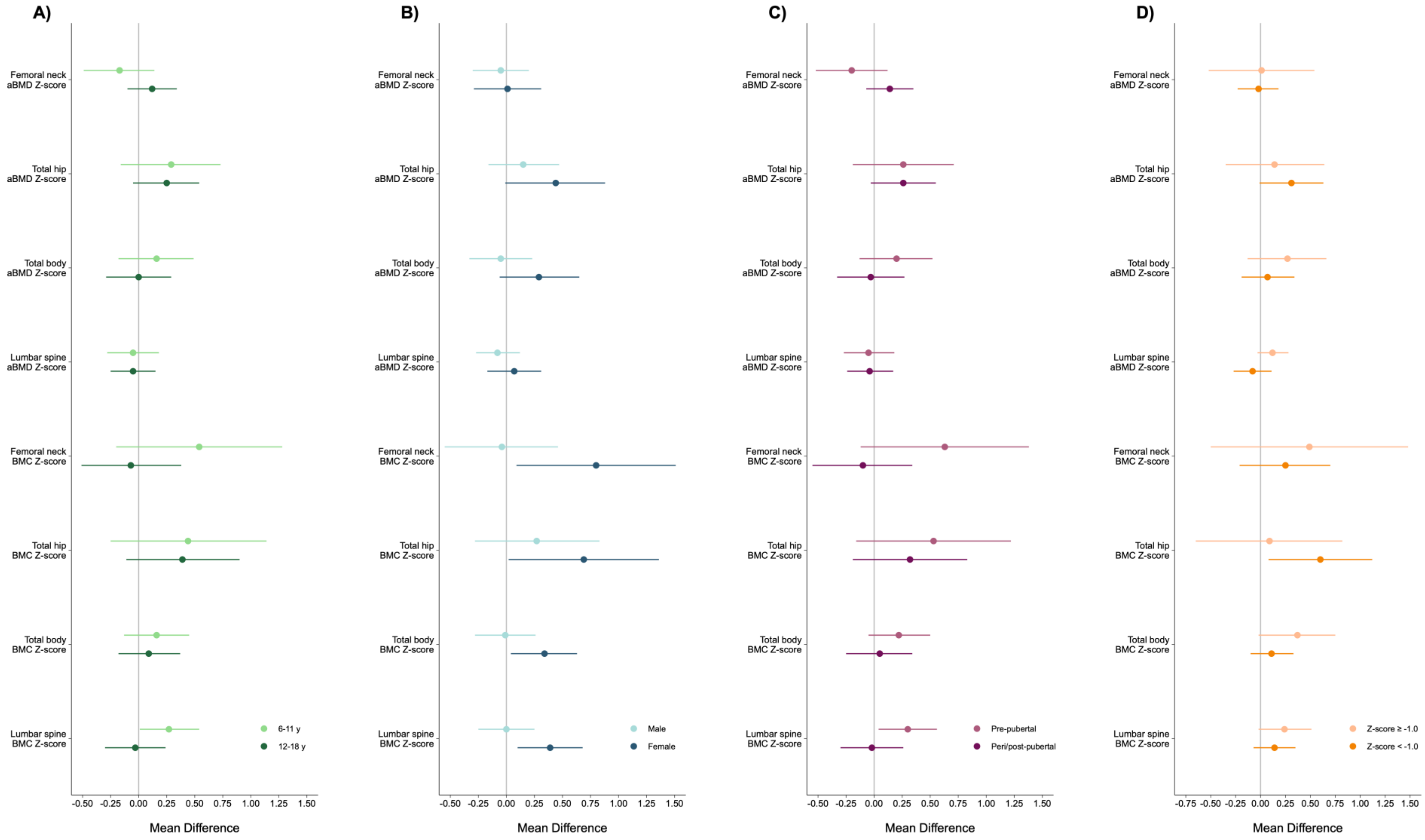
A)
B)
C)
D)
Femoral neck aBMD Z-score
Total hip aBMD Z-score
Total body aBMD Z-score
Lumbar spine aBMD Z-score
Femoral neck BMC Z-score
Total hip BMC Z-score
Total body BMC Z-score
Lumbar spine BMC Z-score
-0.50 -0.25 0.00 0.25 0.50 0.75 1.00 1.25 1.50
-0.75 -0.50 -0.25 0.00 0.25 0.50 0.75 1.00 1.25 1.50
Mean Difference
6-11 y
12-18 y
Male
Female
Pre-pubertal
Peri/post-pubertal
Z-score ≥ -1.0
Z-score < -1.0

**Fig. S1.** Intention-to-treat effects by age (A), sex (B), somatic maturity (C), and low aBMD/BMC at baseline (D). Dots represent the between-groups mean difference and error bars indicate 95% confidence intervals. Data were analyzed using a constrained longitudinal data analysis (cLDA) with linear outcomes, assuming baseline means of the outcome measure were identical between groups. Age-, sex-, and race-specific aBMD and BMC Z-scores at each site are presented using international reference data from the Bone Mineral Density in Childhood Study *(2)*.

Somatic maturity groups were separately created for prepubertal (< -1 year from PHV) and peri/post-pubertal survivors (≥ -1 year from PHV) in accordance with Faigenbaum et al. *(3)*. Similarly, survivors with and without low aBMD/BMC at baseline (Z-score < -1 and Z-score ≥ -1) were stratified. Abbreviations: aBMD, areal bone mineral density; BMC, bone mineral content.

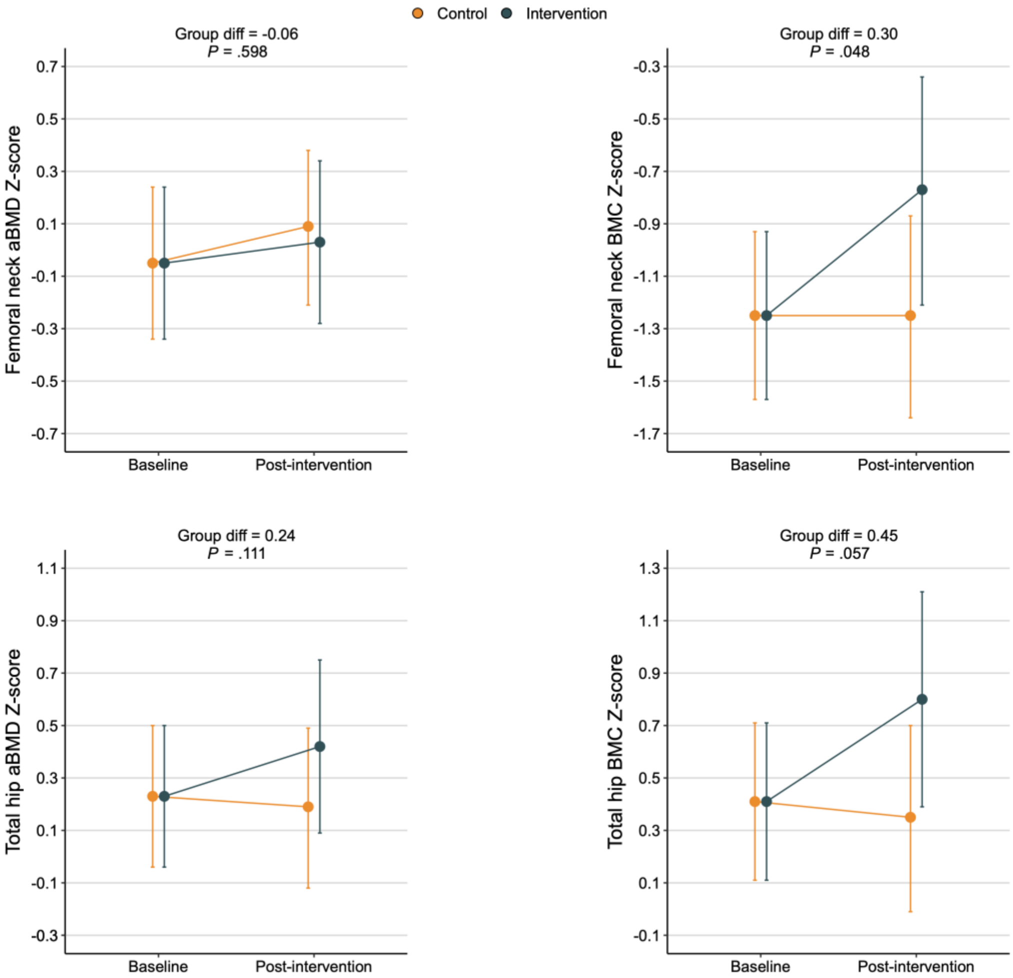


**Fig. S2.** Estimated means at constrained baseline and 9-month post-intervention follow-up in the aBMD and BMC Z-score outcomes. Error bars represent 95% confidence intervals. Data were analyzed using a constrained longitudinal data analysis (cLDA) with linear outcomes, assuming baseline means of the outcome measure were identical between groups (Per-protocol analysis). Abbreviations: aBMD, areal bone mineral density; BMC, bone mineral content; diff, Difference.

3. AD. Faigenbaum, DG. Lloyd, JL. Oliver, *Essentials of youth fitness* (Human Kinetics, Champaign, 2020).
